# Evaluation of Atrial Fibrillation Detection in short-term Photoplethysmography (PPG) signals using artificial intelligence

**DOI:** 10.1101/2023.03.06.23286847

**Authors:** Debjyoti Talukdar, Luis Felipe de Deus, Nikhil Sehgal

## Abstract

Atrial Fibrillation (AFIB) is a common atrial arrhythmia that affects millions of people worldwide. However, most of the time, AFIB is paroxysmal and can pass unnoticed in medical exams therefore regular screening is required. This paper proposes machine learning methods to detect AFIB from short-term ECG and PPG signals. Several experiments were conducted across five different databases with three of them containing ECG signals and the other two consisting of only PPG signals. A total of 269,842 signal segments were analyzed across all datasets (212,266 were of normal sinus rhythm (NSR) and 57,576 corresponded to AFIB segments). Experiments were conducted to investigate the hypothesis that a machine learning model trained to predict AFIB from ECG segments, could be used to predict AFIB from PPG segments. A random forest machine learning algorithm achieved the best accuracy and achieved a 90% accuracy rate on the UMMC dataset (216 samples) and a 97% accuracy rate on the MIMIC-III dataset (2,134 samples). The ability to detect AFIB with significant accuracy using machine learning algorithms from PPG signals, which can be acquired via non-invasive contact or contactless, is a promising step forward toward the goal of achieving large-scale screening for AFIB.

## INTRODUCTION

Atrial fibrillation is an arrhythmia characterized by the absence of the P wave and irregular heartbeats. AFIB is also the most common sustained heart rhythm disorder [1]. Commonly it is associated with deadly and debilitating consequences including heart failure, stroke, poor mental health, reduced quality of life, and death [2]. According to a systematic review published in The Lancet journal, the prevalence of AFIB is estimated to be 46.3 million globally, with 3.8 million new diagnoses annually; this represents an increase of 32% from 2006 to 2016 [3]. Moreover, it has been estimated that 6 to 12 million people will suffer from this condition in the US by 2050 and 17.9 million in Europe by 2060 [4].

There are four different types of AFIB conditions, Paroxysmal AF, Persistent AF, Long-standing Persistent AF, and Permanent AF. However, as stated by [5], only the severest, long-standing Persistent, and Permanent, can be easily detected with an Electrocardiogram exam, while the others are harder to identify due to the irregularity of the symptoms.

These irregularities make the diagnosis of AFIB very difficult, therefore models that can predict AFIB events with short ECG signals have been developed. Those models are usually based on Digital Signal Processing and Artificial Intelligence (AI) techniques. The main goal of these approaches is to make an embedded solution that could be used in a mobile phone, smartwatch, or smart band.

Researchers have put their efforts into two main branches of the Artificial Intelligence field, Machine Learning, and Deep Learning. In the past few years, Deep Learning has become a hot topic in the context of computing and is widely applied in various application areas like health-care, visual recognition, and many more [6]. Convolution Neural Networks (CNN) have been used to detect AFIB [7]; the method proposed by Ross-Howe and Tizhoosh uses ECG signals from MIT-BIH Atrial Fibrillation Database v1.0 [8]. Spectrograms were computed from a six-second ECG window and the CNN was then trained using these spectrograms. This method achieved a reported sensitivity of 98.33%, specificity of 89.74%, and accuracy of 93.16%.

Bruun et al. [9] have used Machine Learning models with a combination of Heart Rate Variability (HRV) and Discrete Wavelet Transform features. ECG signals from the MIT-BIH dataset were segmented using a 30-seconds sliding window with a 5-second stride. Features were extracted from each window and used to create a bootstrap ensemble method with aggregation on decision trees. This method achieved a reported sensitivity of 87.97%, specificity of 96.62%, and accuracy of 93.33%.

There are other authors who have tried to combine the advantages of deep neural networks with handcrafted features. Hu et al. [10] have used the AF Classification from a Short Single Lead ECG Recording: The PhysioNet/Computing in Cardiology Challenge 2017 [11] dataset to extract multiple features. The handcrafted features represent HRV, morphological, frequency domain, and others. A Residual Neural Network (ResNet) was used to extract deep features from a single heartbeat, and after training the ResNet, the in-depth features of the last layer were extracted. Ultimately, a Random Forest model with 1,000 classifiers was trained using the handcrafted and deep features. Reported results show a sensitivity of 88.70%, specificity of 99.60%, and accuracy of 96.30%.

The aim of this study is to develop and compare two different techniques to detect AFIB events in short-term ECG signals as well as verify the feasibility of using PPG signals. The first is based only on signal processing, while the second method uses AI models to predict AFIB.

The remainder of this paper is structured as follows. Section II presents an overview of the methodology, which consists of the details about the datasets used in this work, a full description of the two proposed methods, and the evaluation metrics used. In section III we present the results of the experiments that were conducted. In section IV we discuss our results and the analyses that seek to support the performance achieved. Finally, section V concludes the article by summarizing the findings and discussing future work ideas.

## MATERIALS AND METHODS

### A. Datasets

In this work, five different datasets were used, all of them publicly available. The datasets used are from Physionet’s database [12], named: MIT-BIH Atrial Fibrillation Database v1.0 (AFDB) [8], AF Classification from a Short Single Lead ECG Recording: The PhysioNet/Computing in Cardiology Challenge 2017 (AFC) [11] and MIT-BIH Normal Sinus Rhythm Database (NSRDB), MIMIC III Dataset [13] and UMMC Simband Dataset [14]. Each one of them will be described in the following sections.

#### 1. AFDB

This dataset consists of 23 long-term ECG recordings, each sample has around 10 hours of ECG signals sampled at 250 Hz, with 12-bit resolution over a range of ±10 millivolts. Alongside the ECG signal, there are annotation files that were manually prepared by physicians which contain rhythm annotations of types AFIB (atrial fibrillation), AFL (atrial flutter), J (AV junctional rhythm), and N (normal sinus rhythm).

In this work only Normal and AFIB events were used. Since the dataset has long-term signals with multiple Normal and AFIB events, using the annotation file provided, the signals were divided into events and treated individually. For example, if in a given signal there is Normal sinus rhythm from sample 0 to 10,000 and then AFIB from 10,001 to 20,000 this signal will be divided into two blocks and treated as individual samples.

#### 2. AFC

The 2017 PhysioNet/CinC Challenge dataset was created to encourage the development of algorithms to classify, from a single short-term ECG lead recording, whether the recording shows normal sinus rhythm, atrial fibrillation (AF), an alternative rhythm, or is too noisy to be classified. The available training set contains 8,528 single lead ECG recordings lasting from 9 s to just over 60 s, sampled at 300 Hz.

In this work, only signals classified as Normal or AFIB were used.

#### 3. NSRDB

This database includes 18 long-term ECG recordings of subjects that have had no indication of significant arrhythmia. Data include 5 men, aged 26 to 45, and 13 women, aged 20 to 50. Each sample has about 25 hours of ECG signals sampled at 128 Hz. However, because of the large amount of data, only 2 hours of data were used.

#### 4. MIMIC-III Dataset

The MIMIC-III (Medical Information Mart for Intensive Care) is a large, single-center database comprising information relating to patients admitted to critical care units at a large tertiary care hospital. This database provides continuous ECG and PPG signals from patients in critical care at a large tertiary care hospital [13]. Ten subjects were selected from the MIMIC III database by [14] and segmented in windows of 30 seconds long. In this study, only PPG signals were used, which were downsampled to 50 Hz.

#### 5. UMMC Simband Dataset

The UMMC dataset [14] consists of 37 patients (28 male and 9 female) with cardiac arrhythmia, aged between 50 and 91 years old. The participants were asked to wear the smartwatch Simband 2 (Samsung Digital Health, San Jose, CA, USA) and had their ECG reference taken using a 7-lead Holter monitor (Rozinn RZ153+ Series, Rozinn Electronics Inc., Glendale, NY, USA). Data were preprocessed and segmented in 30-second windows with no overlapping; ECG data was sampled at 128 Hz while PPG data were downsampled to 50 Hz. All the signals were labeled within 5 categories: 0-Normal Sinus Rhythm (NSR), 1-Atrial Fibrillation (AF), 2-Premature atrial contractions/Premature ventricular contractions (PAC/PVC), 3-Not sure if it is NSR or PAC/PVC, 5-Noisy PPG, NaN-No ECG Reference. However, only PPG signals were used in this study, separated into two labels, AF (1) and Normal (0,2,3); labels 5 and NaN were not used.

### B. Preprocessing

Firstly, the raw ECG signal is obtained from the datasets. The raw ECG is filtered using a smoothing filter Savitzky Golay [15] of 4th order and 19 frames. The smoothed ECG signal is then filtered using a band-pass filter, a 2nd order Butterworth filter [16] with cutoff frequencies of 0.5 Hz - 20 Hz. Ultimately, the signal is normalized using the MinMax approach as described in Eq.1.

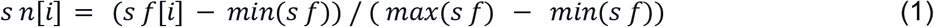

- s n: Signal Normalised
- s f: Signal Filtered

Once the ECG signals were processed, the peaks of the R-wave were extracted. A custom peak detector algorithm was created alongside a double peak correction routine to cope with erroneous peaks.

The third step was to create the Inter-beat signal, which describes, in terms of samples, the distance in time between each peak. The signal as shown in Figure 1 is normalized to the same length as the ECG signal, so each ECG sample has the correspondent inter-beat-interval (IBI).

**Figure 1:**
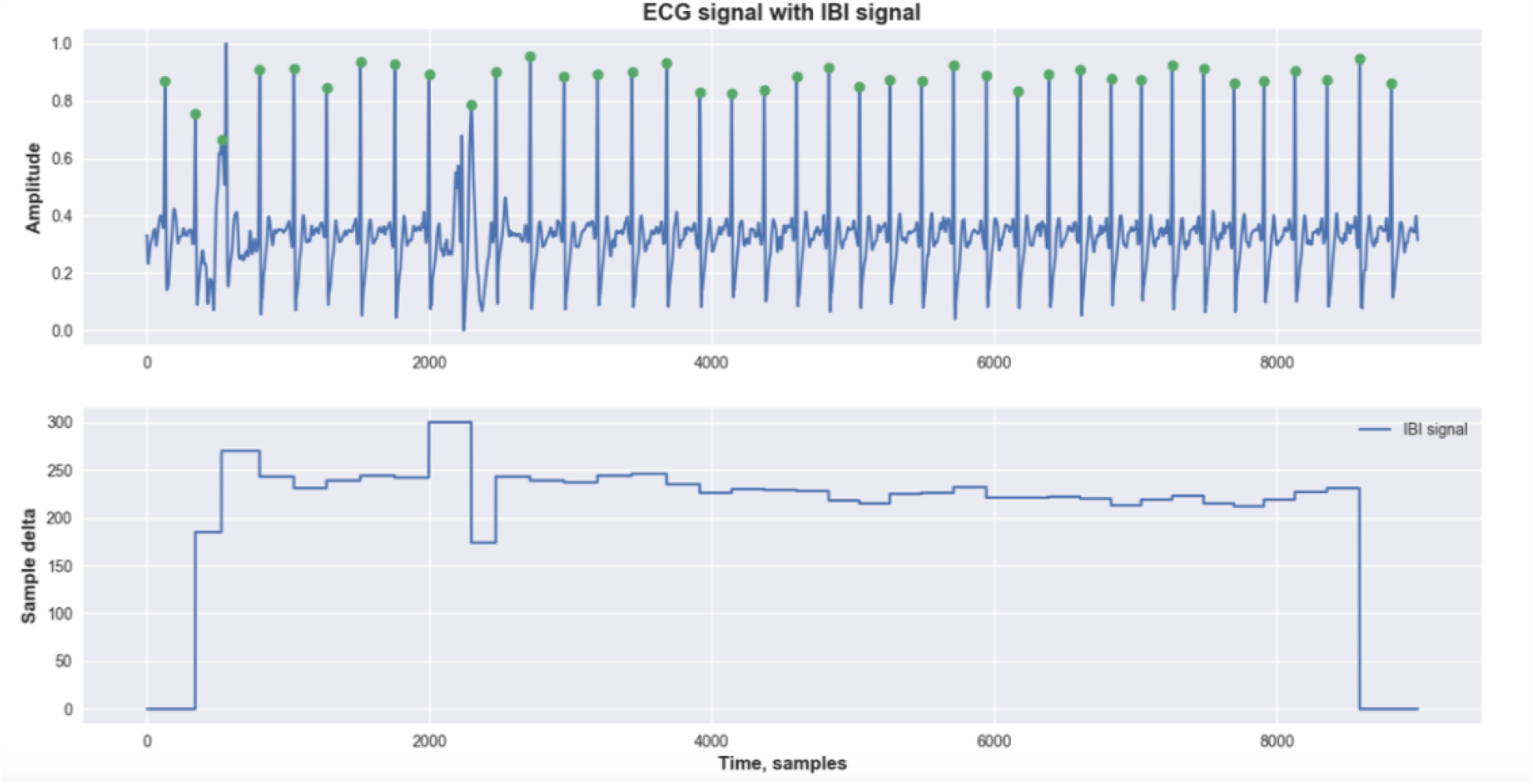
Example of preprocessing layer - ECG signal with highlighted peaks and IBI signal.

The IBI signal carries information about changes in the cardiac rhythm. However, it is well known that signals from public databases are not always perfect and may be noisy, which can lead to wrong predictions. Thus, we propose another routine to validate or invalidate parts of the signal based on the IBI signal. This routine works as a mask; where there are validated IBI segments, we keep the inter-beat-interval, however, where there are invalidated IBI segments, we prune the interval. Eq.2 describes the conditions outlined to invalidate or keep corresponding IBI intervals.

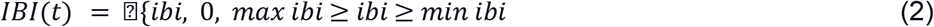

Once we obtained the validated IBI signal, a sliding window approach was used to search for reliable frames within the IBI signal. The frame is considered reliable if in a given window size there were not any invalidated segments, which means that those frames have good signal quality.

### C. Digital Signal Processing Method

The first method uses only signal processing techniques to identify AFIB events. In fact, most of the method’s functions were already described in section II B.

After analyzing the IBI signal, as stated previously, frames of the ECG signal were extracted with an 8-second sliding window along with a stride of 2 seconds. The method will analyze frame by frame and extract a new IBI signal for each one. The first derivative of each frame’s IBI signal was computed. The first derivative shows significant spikes where there were sudden changes in the IBI signal, thus, using a simple threshold it is possible to identify abnormal changes in the cardiac rhythm.

For each frame, two parameters are extracted. The first was a Boolean that represents if this frame has at least one abnormal change and the second was a float number that represents the ratio of abnormal changes, which was calculated by using the ratio between abnormal spikes in the first derivative of the IBI signal and the number of peaks within this frame.

Ultimately, this signal will be classified as AFIB if the mean ratio of abnormal changes between all frames is greater than a threshold.

### D. Machine Learning Method

Secondly, as stated previously, the most common approach to detect AFIB events in short-term signals is by using AI techniques. Therefore, this work proposes its own AI model to predict AFIB.

The method uses the same preprocessing layer as stated previously in section II B.

For each frame, instead of looking at the first derivative of the IBI signal, this method seeks to extract HRV features in a given window. This work has used a sliding window approach with 15 seconds in size and a 2-second stride.

The HRV features were used as an input to the ML models, and this work has evaluated the performance of three approaches: Random Forest (RF), Support Vector Machine (SVM), and K-Nearest Neighbours (KNN).

Seven different HRV features were selected based on Permutation Features Importance:

- HRMAD: Mean Absolute Deviation of Heart Rate.
- RMSSD: Root Mean Square of Successive Differences between Normal Heartbeats.
- IRQNN: Inter-Quartile Range of Normal Heartbeats.
- MCVNN: HRMAD divided by the median of Normal Heartbeats.
- CVNN: Standard Deviation of Normal Heartbeats divided by the mean of Normal Heartbeats.
- CVSD: RMSSD divided by the mean of Normal Heartbeats.
- HTI: HRV Triangular Index.

The HRV features were extracted per frame and each entry represents one row with seven features and the correspondent label, AFIB or Normal.

However, given the sliding window approach, we may end up with multiple entries for the same individual so it is essential to ensure the same individual is not included in both the training and test dataset.

The dataset with HRV features was divided into two groups as follows: 60% of the individuals in the training set and 40% in the testing set. It is worth mentioning that once there are a different number of entries per individual and therefore 60% of the individuals do not represent 60% of the available data.

Finally, Figure 2 (a) and (b) show the respective block diagrams of the proposed approach.

**Figure 2 (a).**
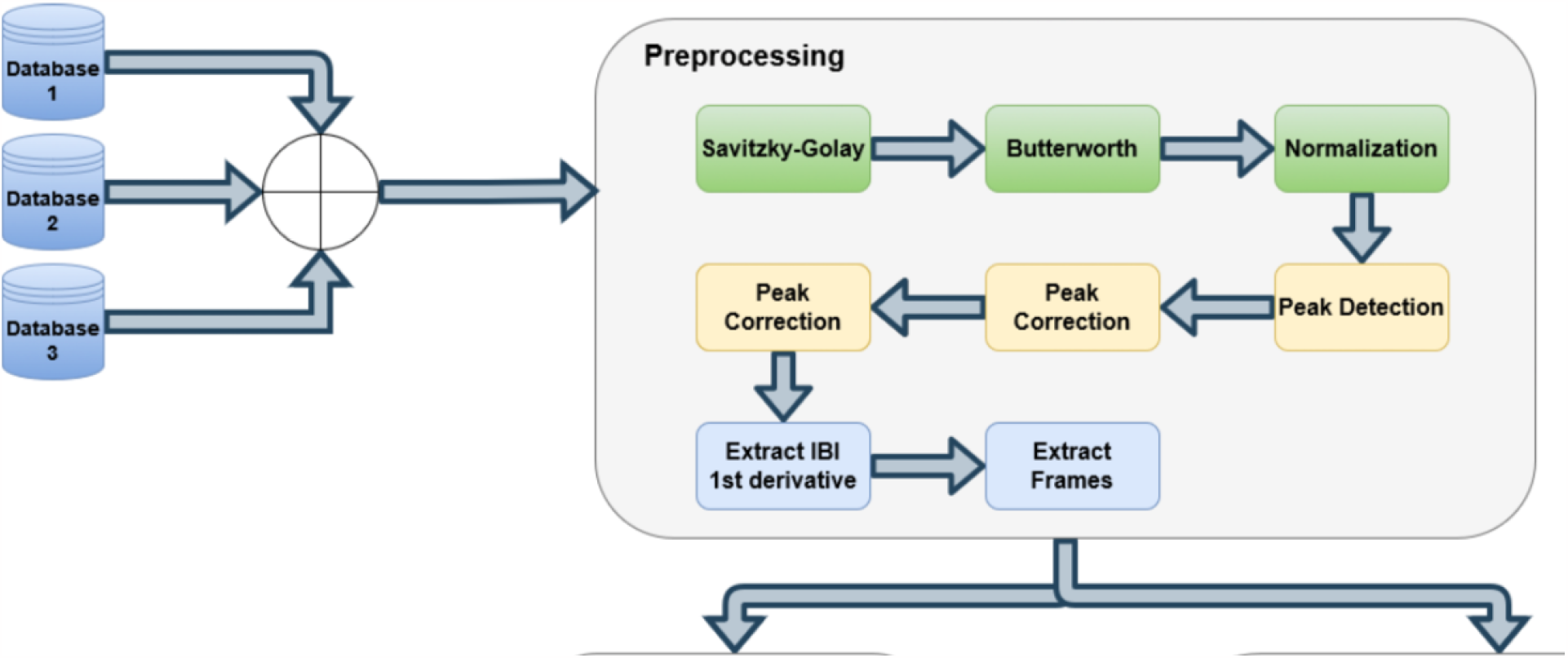
Block diagram part 1

**Figure 2 (b).**
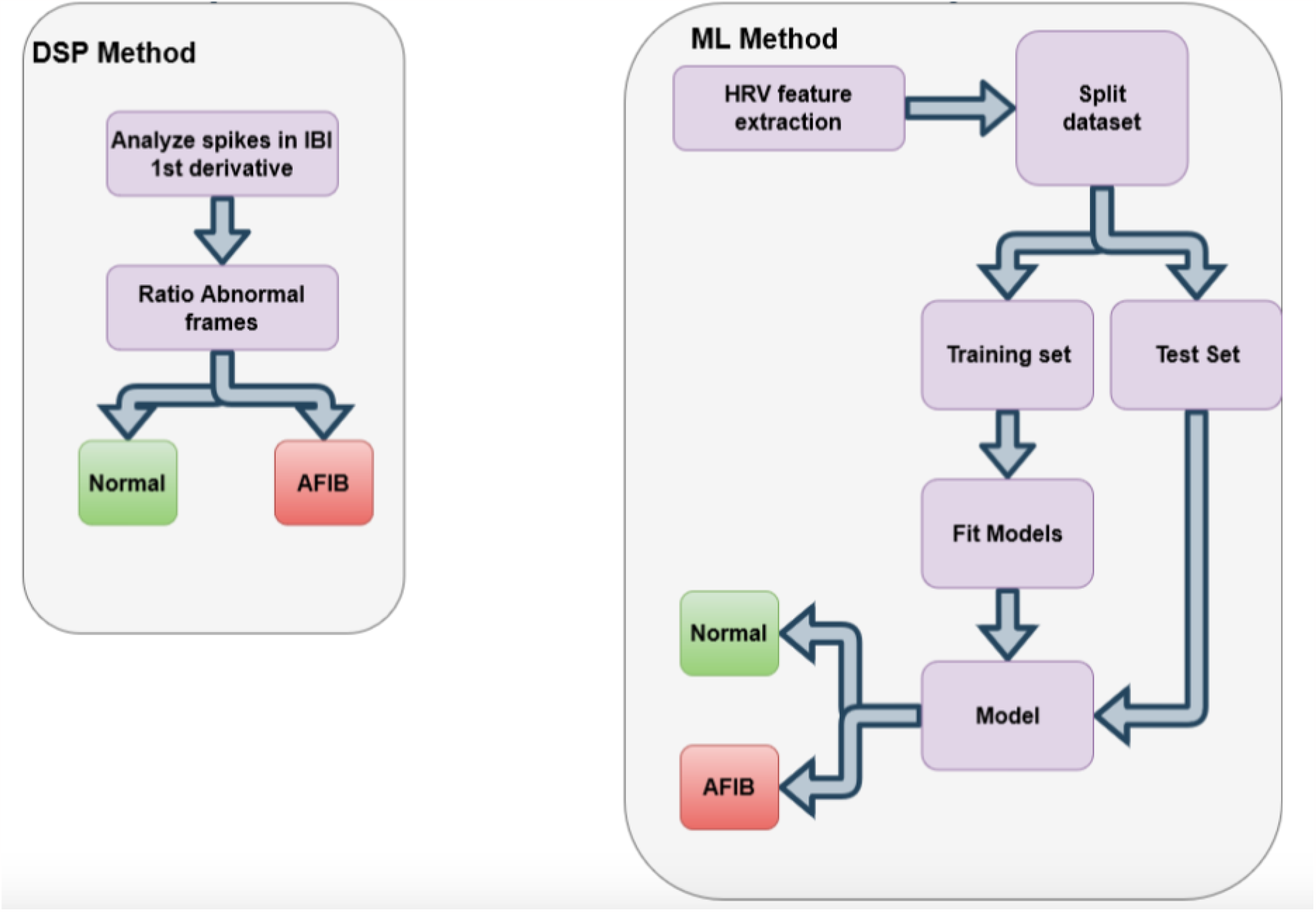
Block diagram part 2

### E. Metrics

In order to evaluate the performance of the proposed methods, this work chose to use a set of objective metrics that will reflect the model’s performance. All the metrics, described in Eqs.3,4,5,6 are based on the following concepts:

- True Positive (TP): Event in which there was AFIB and it was correctly identified.
- False Positive (FP): Event in which there was no AFIB and the algorithm pointed it incorrectly as AFIB.
- True Negative (TN): Event in which there was no AFIB and the algorithm pointed it correctly as Non-AFIB.
- False Negative (FN): Event in which there was AFIB and the algorithm has not found it.

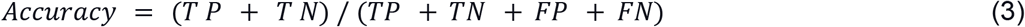

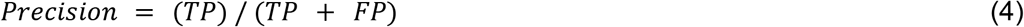

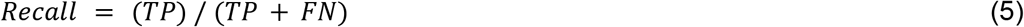

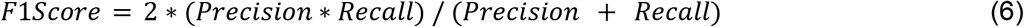

## RESULTS

Figures 3 and 4 show examples of the DSP method. In both figures, the top plot represents the ECG signal in blue with the highlighted peaks and the red marks refer to the portion of the signal that was analyzed in each frame. In the bottom plot, the IBI signal is shown in blue and its first derivative is shown in green; the red line represents the threshold for abnormal spikes. Figure3 shows an example of a Normal ECG while Figure 4 an ECG with AFIB.

**Figure 3:**
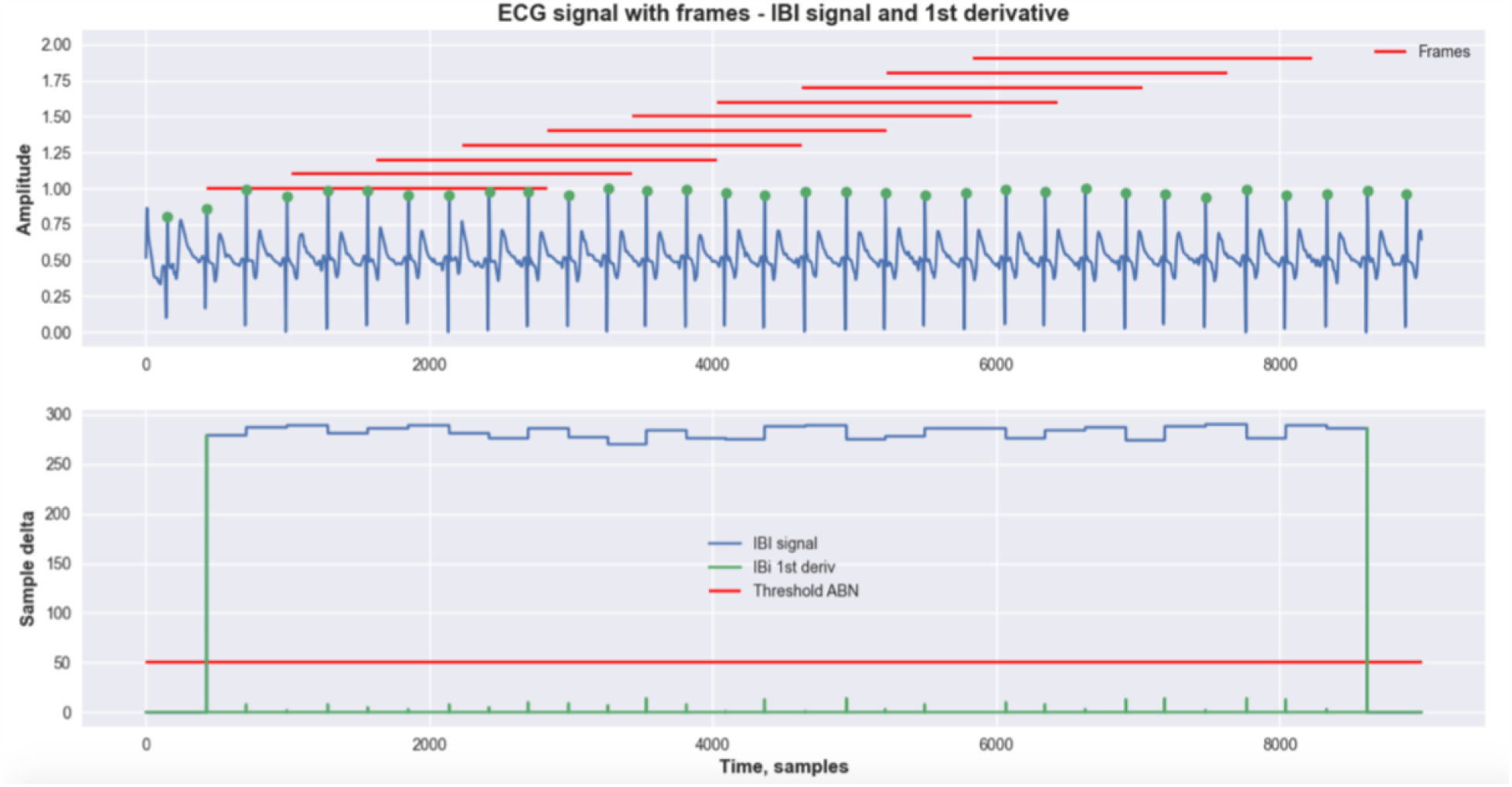
Example of execution of DSP method - Normal ECG.

**Figure 4:**
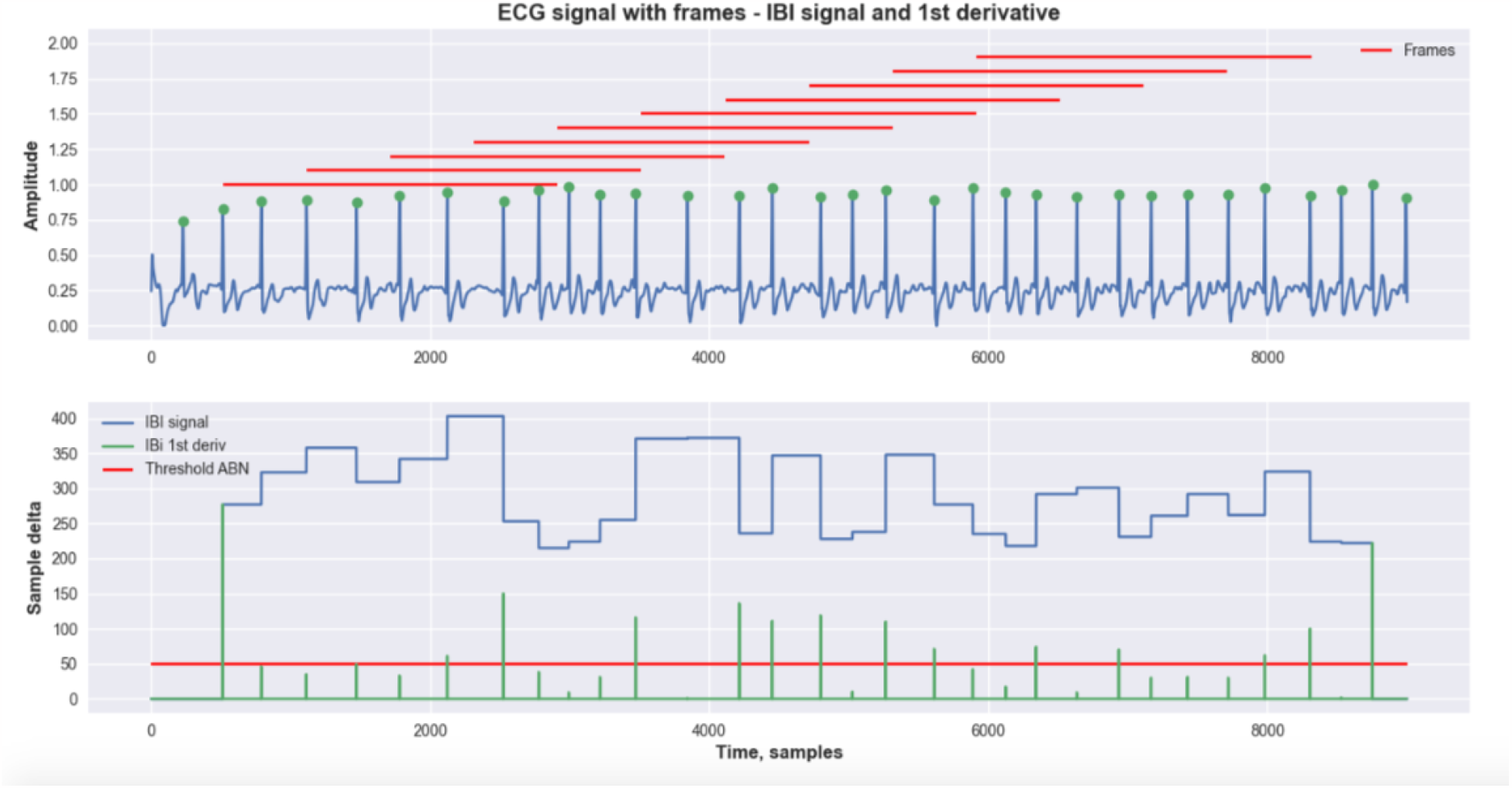
Example of execution of DSP method - AFIB ECG.

One of the advantages of this method is that it does not require any training process, thus, it is possible to use all the available data as a test set.

Figure 4: Example of execution of DSP method - AFIB ECG.

Furthermore, all three datasets were used individually as separate experiments. Figure 5 shows the class distribution for each dataset.

**Figure 5:**
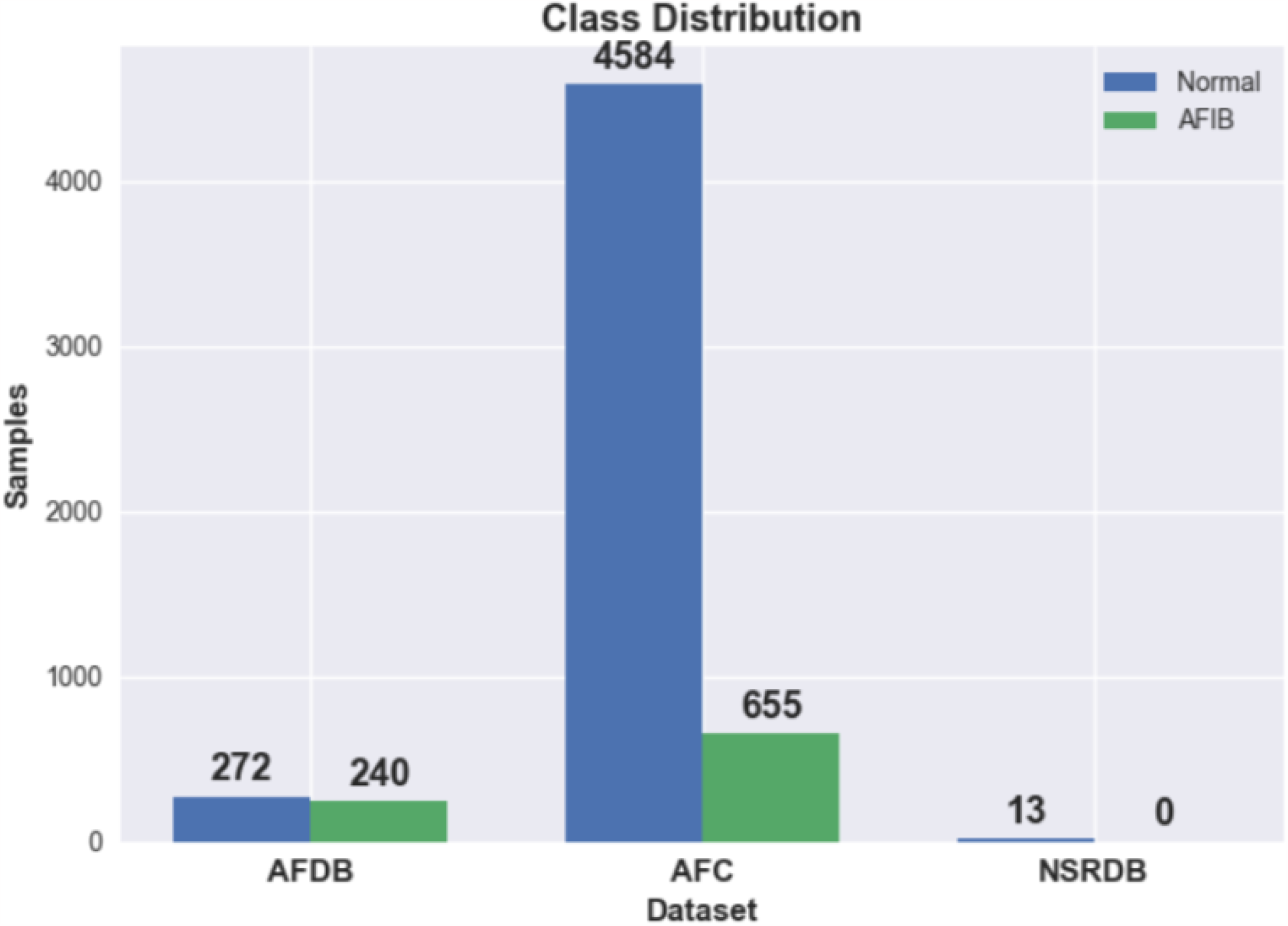
Class distribution in each dataset.

### A. Experiment 1 - AFDB

In the first experiment, data from the AFDB dataset was used. Originally, there were 23 long-term ECG samples, however, as stated previously, each signal was divided into blocks using the annotated events. Consequently, the number of samples increased to 512.

Figure6 shows the confusion matrix for the first experiment using the DSP method. In the first dataset, using the proposed metrics, the method was able to achieve 90.82% of accuracy, 90.98% of precision, 91.01% of recall, and an F1-score of 90.81%.

**Figure 6:**
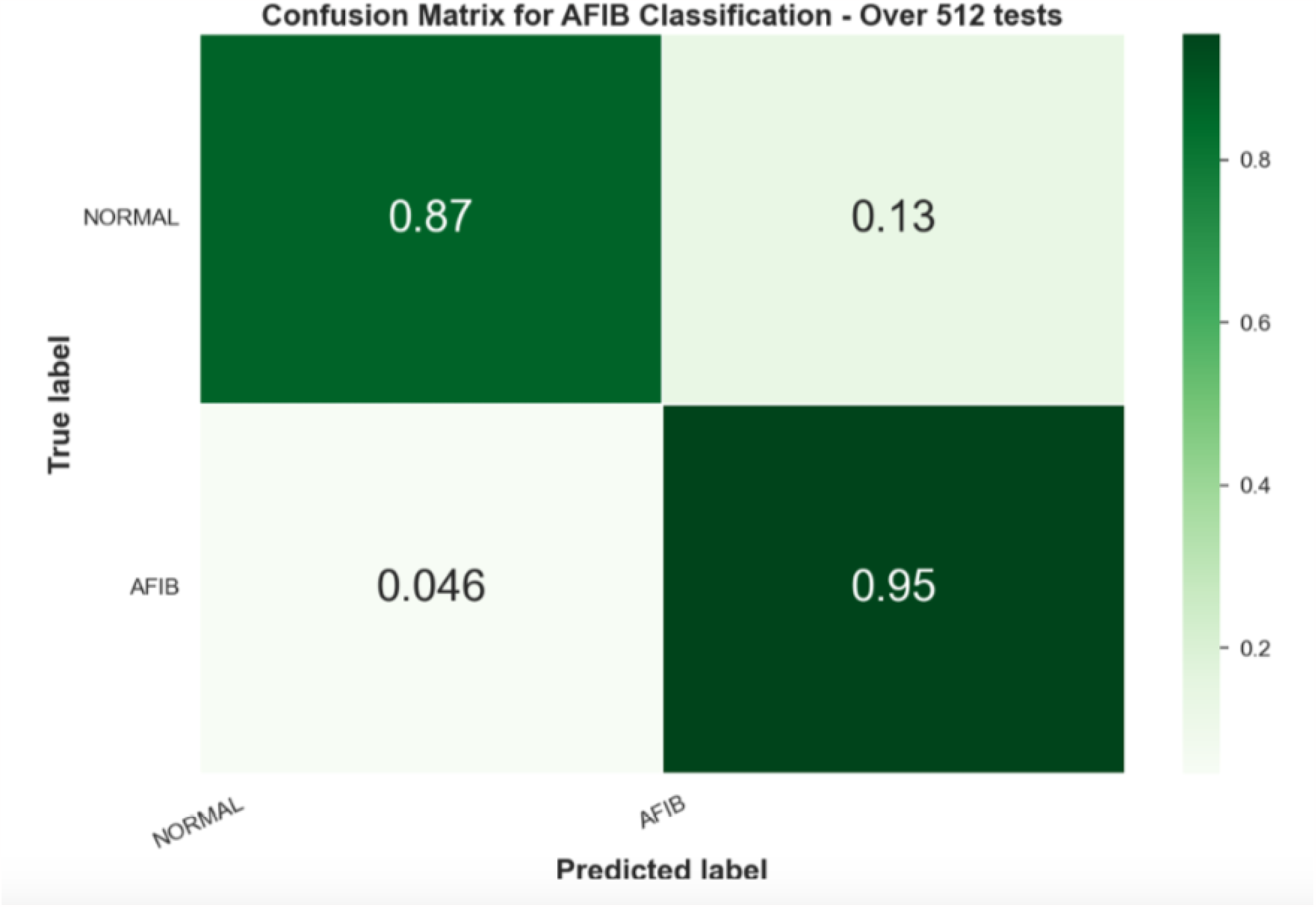
Confusion Matrix for DSP method applied to AFDB dataset.

### B. Experiment 2 - AFC

Secondly, using the AFC dataset, around 5,200 samples survived the preprocessing layer and were analyzed. Figure 7 shows the confusion matrix for the second experiment using the DSP method. In this experiment, the method was able to achieve 87.32% accuracy, 73.59% precision, 86.28% recall, and F1-Score 77.49%.

**Figure 7:**
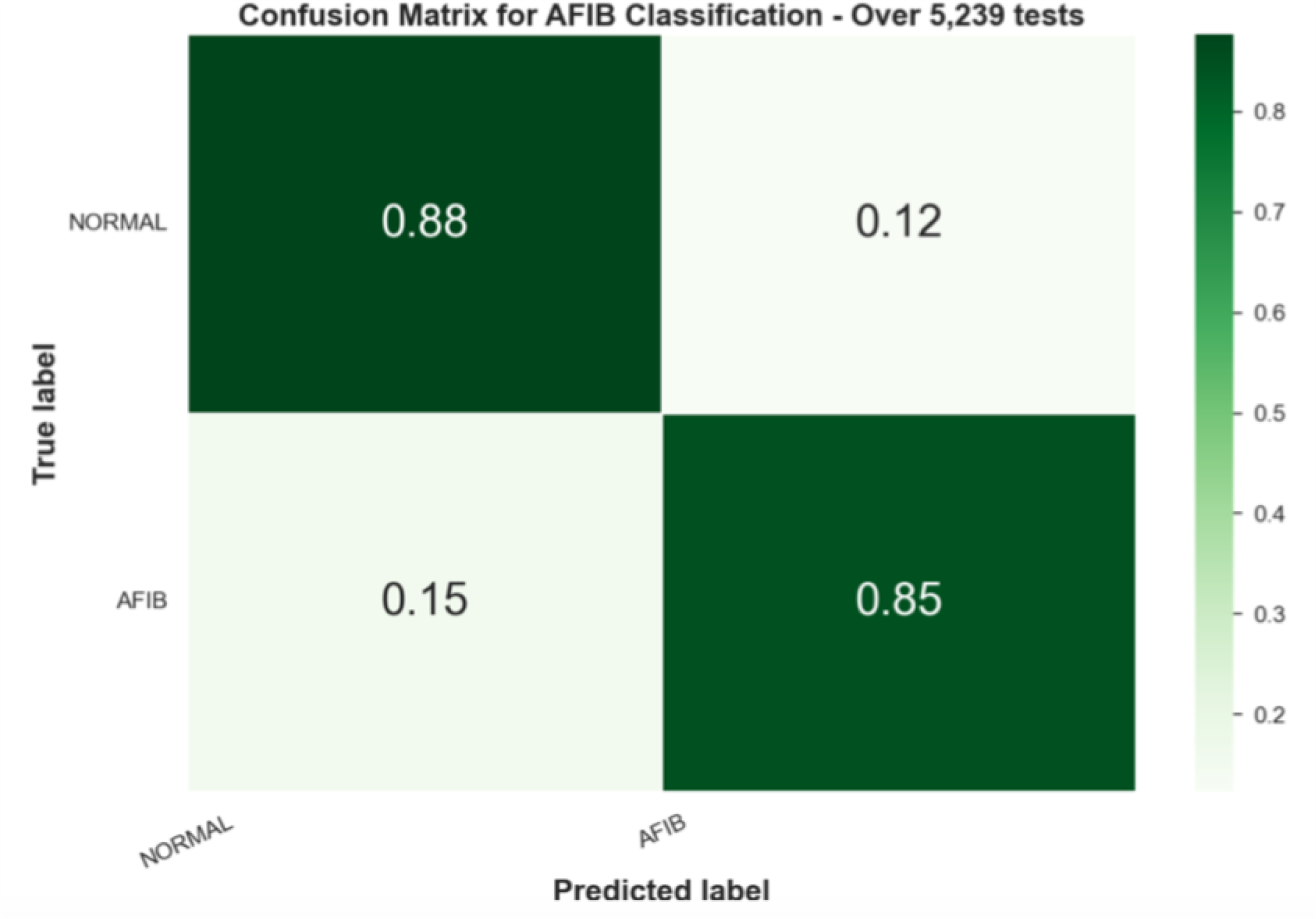
Confusion Matrix for DSP method applied to AFC dataset.

### C. Experiment 3 - NSRDB

Finally, in the last experiment of the first method, the NSRDB dataset was used to evaluate the false positive rate. Thus, the purpose of this experiment was to see how many times the model would incorrectly classify segments as AFIB given there were only normal signals.

Since the NSRDB only has normal samples, neither a confusion matrix nor statistical results would be the best way to show the results so we show the number of correct predictions, in 12 out of 13 samples, resulting in 92.31% accuracy.

In the machine learning approach, HRV features were extracted from each frame of the signal. Hence, the resulting HRV dataframe has multiple entries for one single sample, which means that the dataset class distribution also changes. Figure8 shows the class distribution for the HRV datasets.

**Figure 8:**
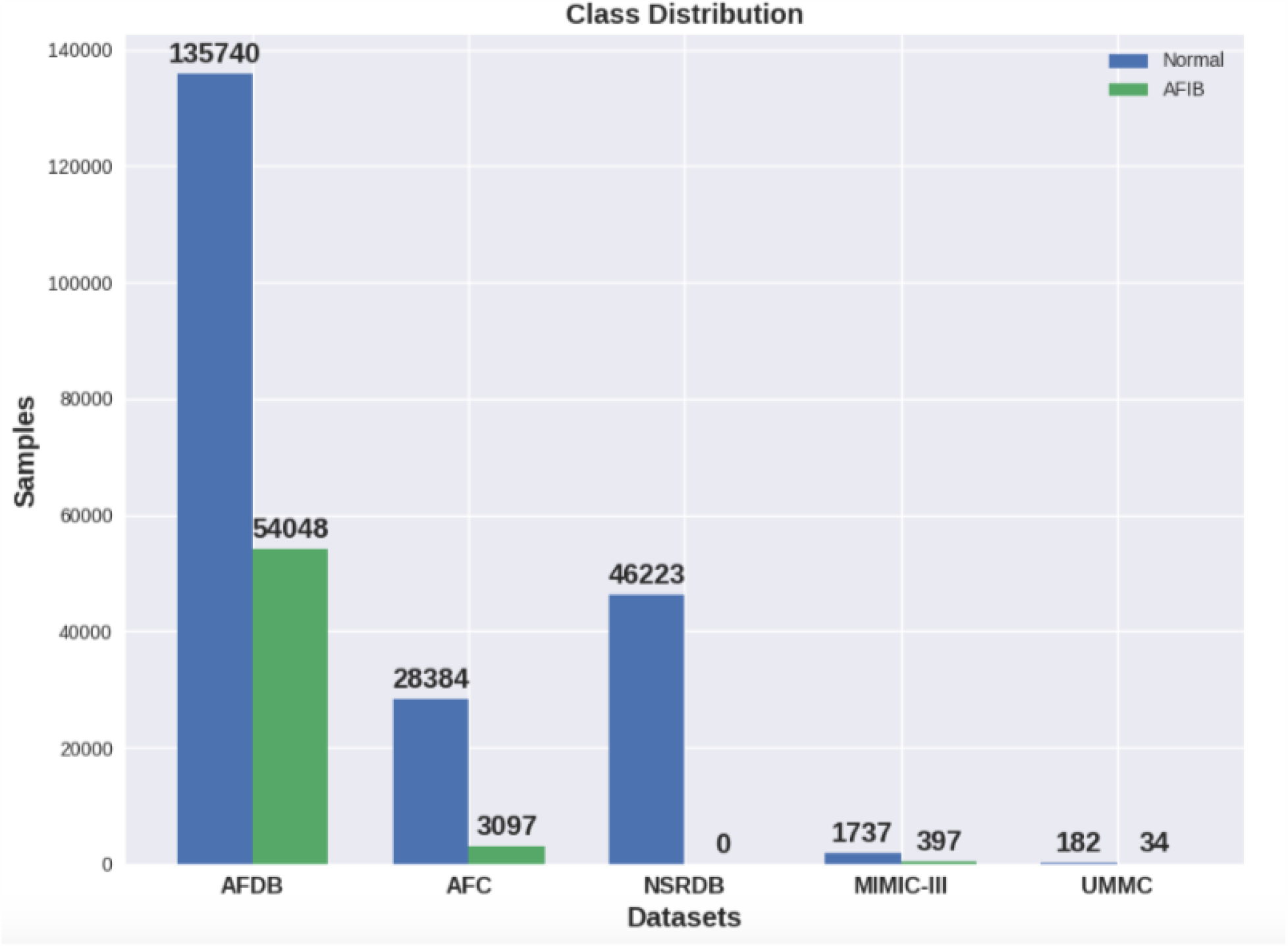
Class distribution in each dataset.

Unlike the DSP method, the machine learning does require a training process, which was carried out with 60% of the individuals from one of the datasets.

### D. Experiment 4 - AFDB testing set

In the first experiment of the ML method, the remaining 40% of the AFDB data, which was not used to train the models, was used as the test set.

Figure 9 Confusion Matrix for ML method applied to AFDB testing set.

**Figure 9.**
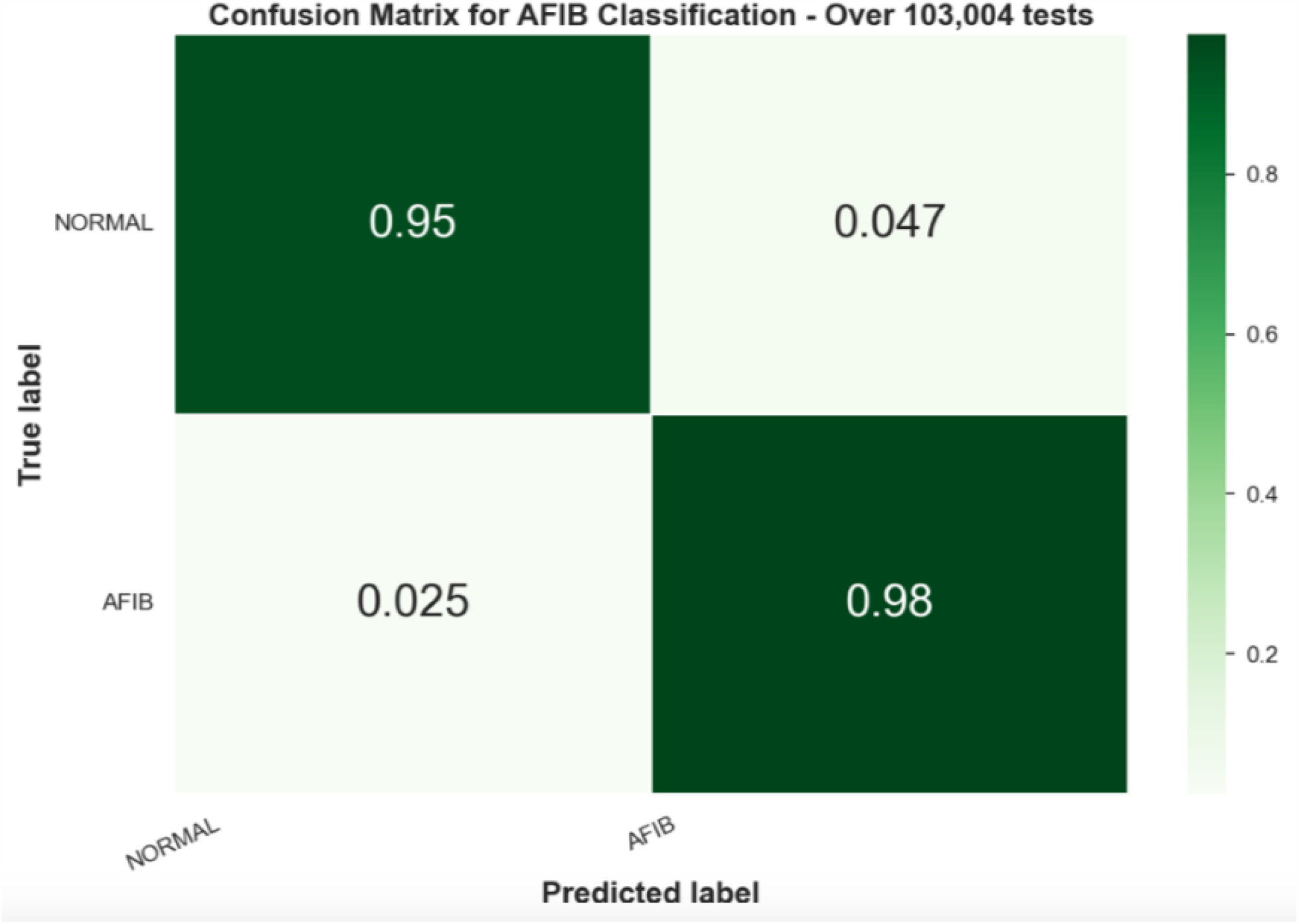
shows the confusion matrix for the fourth experiment with the RF model.

However, this experiment evaluates each HRV entry individually, which means that the model is predicting a label for each frame. Therefore, it is also possible to put all the frames’ predictions together and choose the final prediction of the sample based on a voting system. Figure 10 shows a pie chart with correct and incorrect predictions, in terms of percentage and samples, for Normal and AFIB samples on the AFDB test set.

**Figure 10:**
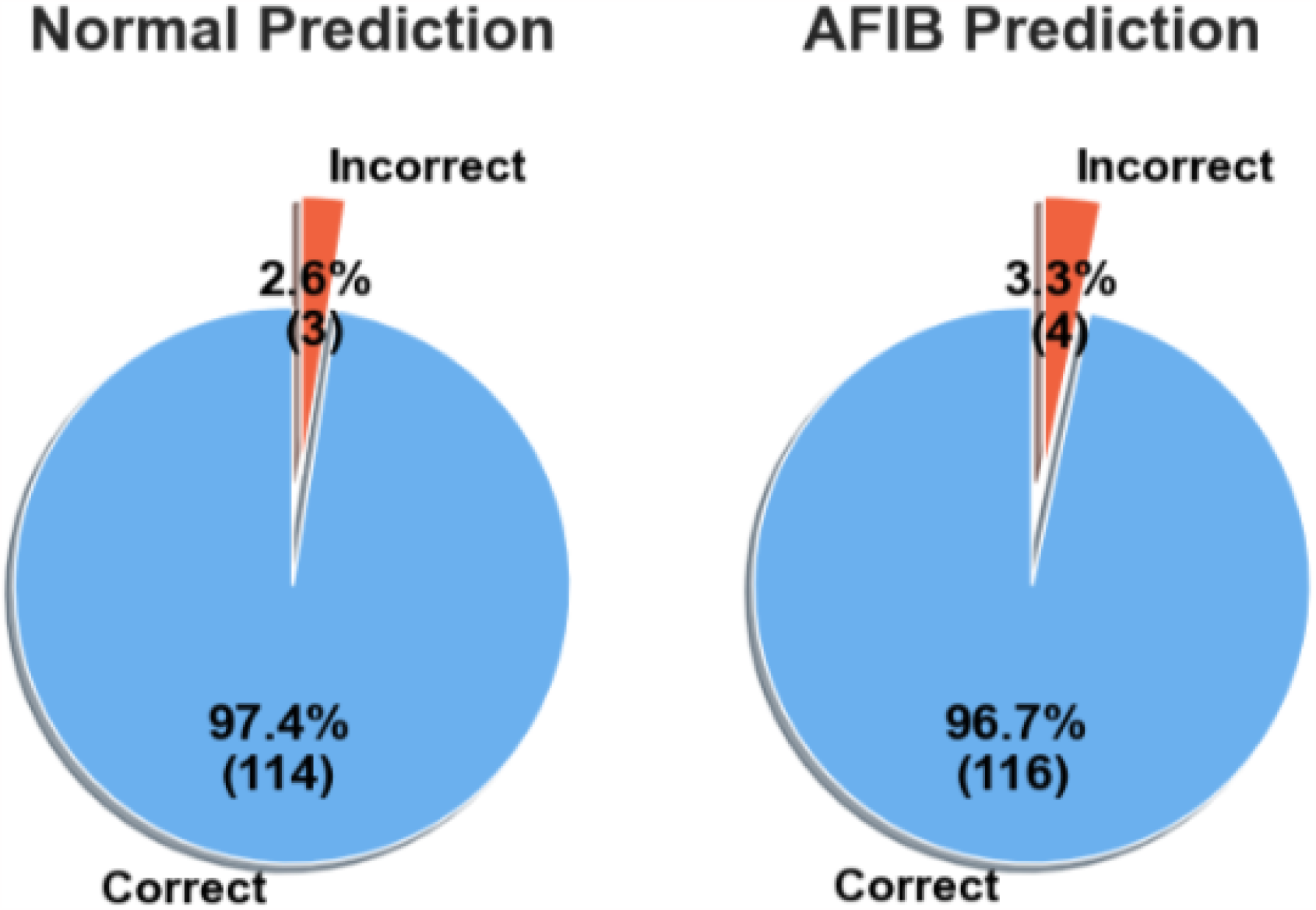
Correct and Incorrect predictions of Normal and AFIB samples on the AFDB testing set.

### E. Experiment 5 - AFC

In the second experiment, the models trained on the AFDB dataset were tested on all the data from the AFC dataset. TableIV shows the statistical performance of each model. In this experiment, the SVM model achieved the best performance with 94.31% accuracy, 83.24% precision, 86.30% recall, and an F1-Score of 84.68%. However, even though the SVM model achieved a slightly better performance, this work chose to keep the analysis with the RF model due to the preference for ensemble methods.

Similar to the previous experiment, it is possible to evaluate the results by choosing the final prediction of the sample using a voting system within the frames’ predictions. Figure 12 shows a pie chart with correct and incorrect predictions, in terms of percentage and samples, for Normal and AFIB samples on the AFC dataset.

**Figure 11.**
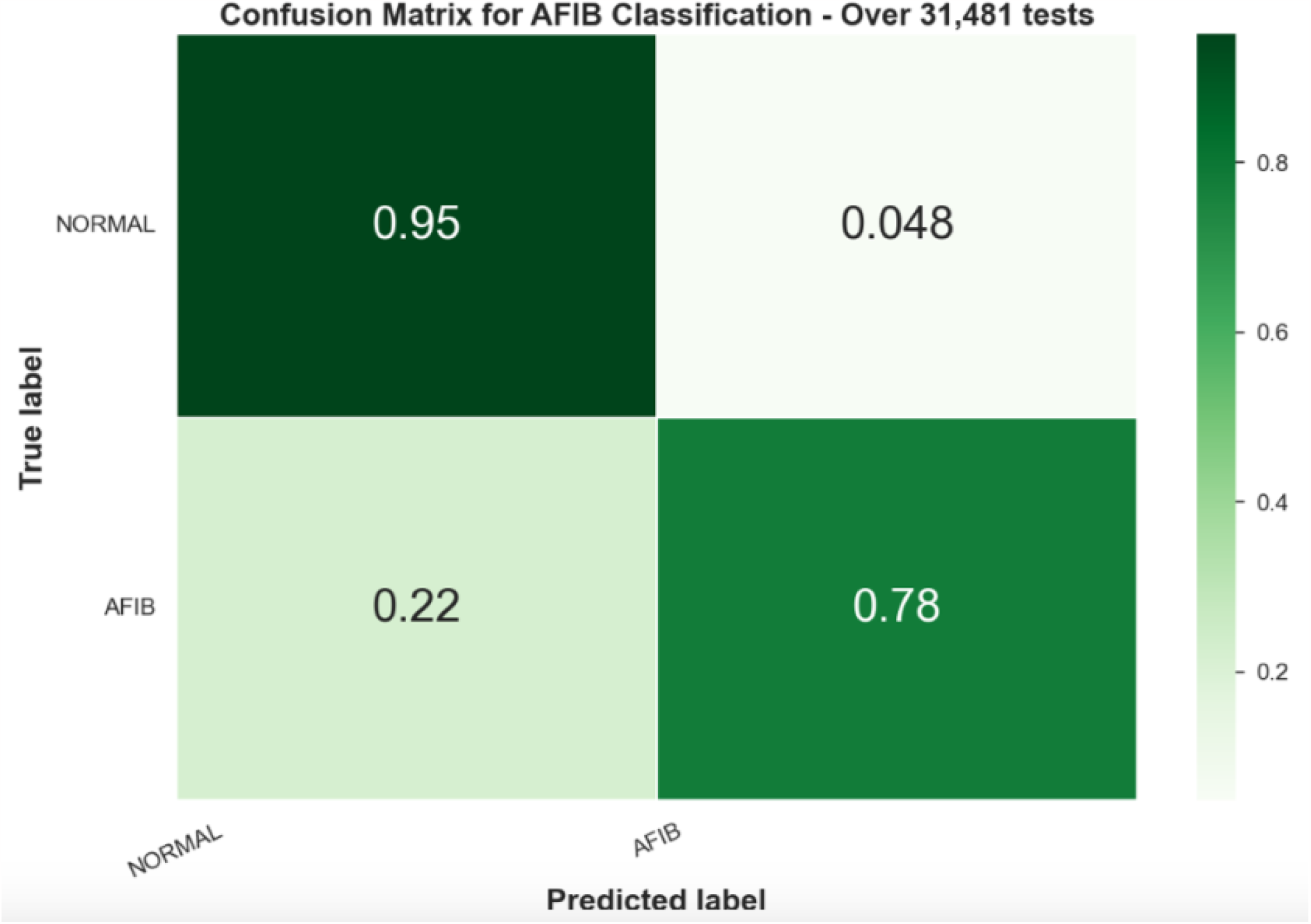
shows the confusion matrix for the fifth experiment with the RF model.

**Figure 12:**
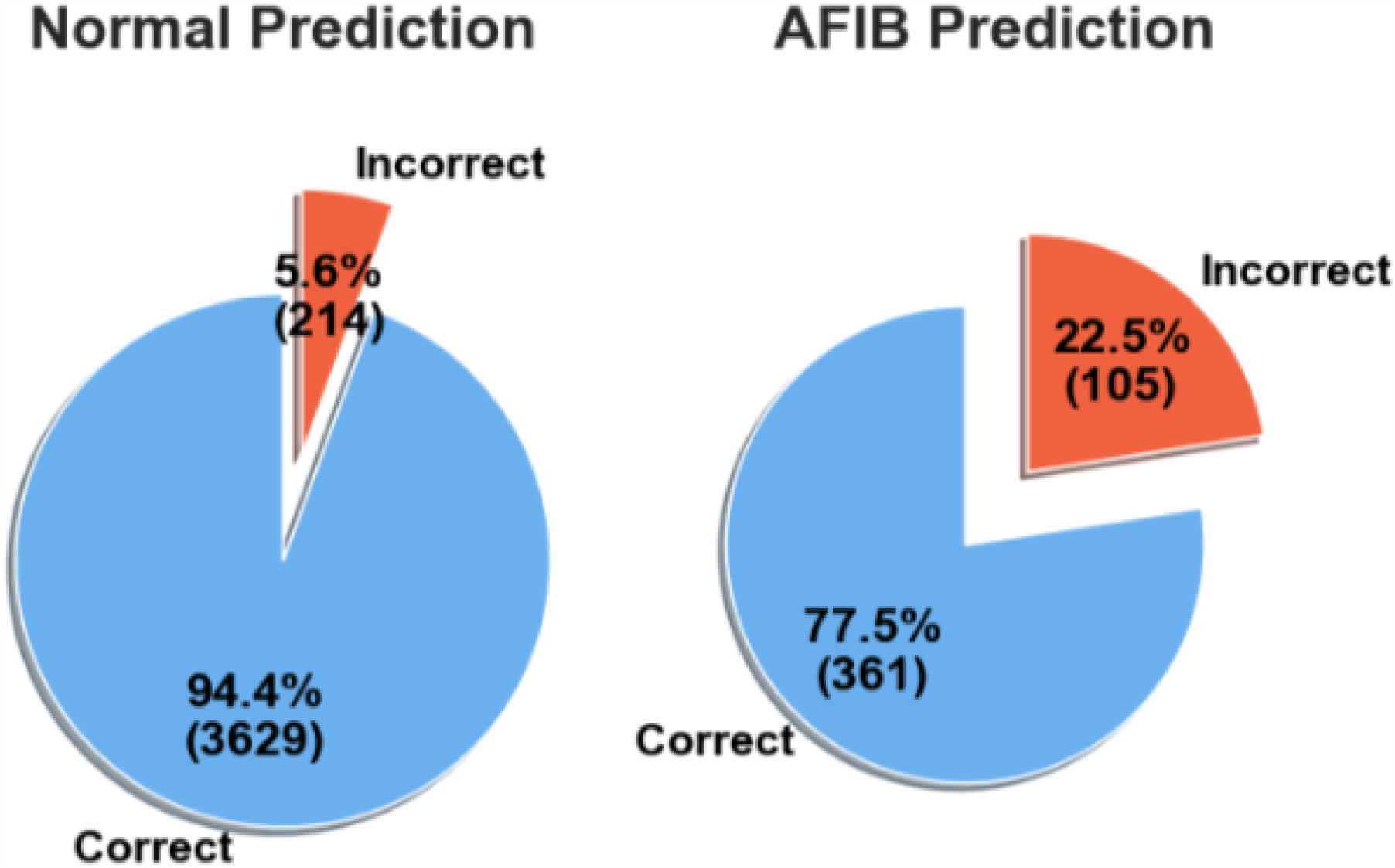
Correct and Incorrect predictions of Normal and AFIB samples on the AFC dataset.

### F. Experiment 6 - NSRDB

The last experiment seeks to evaluate the false positive rate. One of the challenges when using ML models is to make it robust enough to false positives. As stated previously, the NSRDB only has Normal samples, thus, the model was able to correctly predict 41,703 out of 44,399 HRV frames, resulting in 93.93% of accuracy.

When grouping all the frames from respective samples, the model was able to predict correctly as Normal 13 out of 14 samples, resulting in 92.86% of accuracy.

Moreover, Table 4 shows the summary of the statistical results of the ML method when applied to three different datasets with HRV frames.

**Table 1.** shows the statistical performance of each model. As can be seen in Table 1 the Random Forest model achieved the best performance with 96.03% accuracy, 95.08% precision, 96.38% recall, and an F1-Score of 95.67%.

| Dataset | Acc. % | Prec. % | Recall % | F1-Score % |
| --- | --- | --- | --- | --- |
| AFDB | 90.82 | 90.98 | 91.09 | 90.81 |
| AFC | 87.32 | 73.59 | 86.28 | 77.49 |
| NSRDB | 92.3 | - | - | - |

**Table 2:**
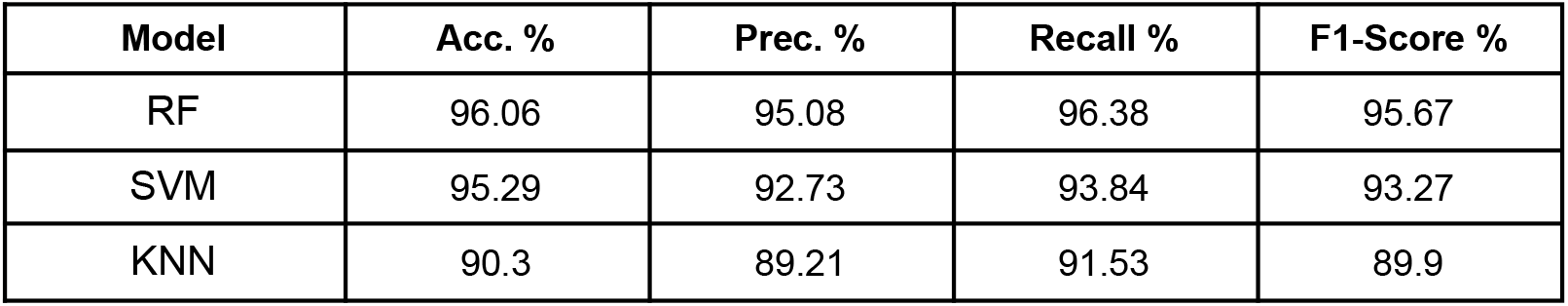
Statistical results for all three models in the ML method with AFDB testing set.

| Model | Acc. % | Prec. % | Recall % | F1-Score % |
| --- | --- | --- | --- | --- |
| RF | 96.06 | 95.08 | 96.38 | 95.67 |
| SVM | 95.29 | 92.73 | 93.84 | 93.27 |
| KNN | 90.3 | 89.21 | 91.53 | 89.9 |

**Table 3:** Statistical results for all three models in the ML method with the AFC dataset.

| Model | Acc. % | Prec. % | Recall % | F1-Score % |
| --- | --- | --- | --- | --- |
| RF | 93.46 | 80.62 | 86.56 | 83.23 |
| SVM | 94.31 | 83.24 | 86.3 | 84.68 |
| KNN | 92.08 | 77.32 | 85.08 | 80.5 |

**Table 4:** Statistical results for the ML method in all three datasets.

| Dataset | Acc. % | Prec. % | Recall % | F1-Score % |
| --- | --- | --- | --- | --- |
| AFDB | 96.06 | 95.08 | 96.38 | 95.67 |
| AFC | 93.46 | 80.62 | 86.56 | 83.23 |
| NSRDB | 93.93 | - | - | - |

Once the ECG tests were conducted, we investigated whether our methods could be applied to PPG signals or not. Even though ECG-based solutions can bring a massive improvement to healthcare areas in detecting AFIB, those still rely mostly on electrodes to properly extract the signal. Thus, a PPG-based solution could be embedded in small devices and be used at home on a daily basis. In our study, we propose an evaluation of our methods applied to two different datasets, a subset of the MIMIC-III dataset and the UMMC Simband dataset.

At this point, based on previous results and analysis, we chose the Random Forest ML method trained using the AFDB dataset as the final solution. The results presented in the next two sessions will describe the performance of the trained ML solution using only PPG signals as the test set.

In this solution two signal quality layers were added in an attempt to prevent poor signals from being predicted, leading to wrong predictions. The first layer is based on the previous method, which uses the IBI signal to detect invalid peaks. Hence, the signal must have at least 75% of the valid peaks to pass this test. Moreover, the Signal Noise Ratio (SNR) was used as an additional layer to prevent making predictions on poor-quality signals. In this work, the signal must have a mean SNR greater than 5 dB to be considered a valid signal.

### G. Experiment 7 - MIMIC-III

In the first PPG experiment, we segmented the PPG signals from the MIMIC-III dataset into 30-second samples. Then, the two-step signal quality check was used to remove bad-quality signal segments. HRV features were extracted from the remaining signal segments that passed the two-step quality check. Figure 8 shows the dataset distribution.

The features extracted were used as inputs into the trained model. The results were collected and presented in Table 5 with statistical metrics as well as the confusion matrix in Figure 13.

**Table 5:** Statistical results for RF ML model trained using AFDB dataset and tested on MIMIC-III dataset.

| Model | Acc. % | Prec. % | Recall % | F1-Score % |
| --- | --- | --- | --- | --- |
| RF | 97.32 | 96.68 | 94.37 | 95.47 |

**Figure 13:**
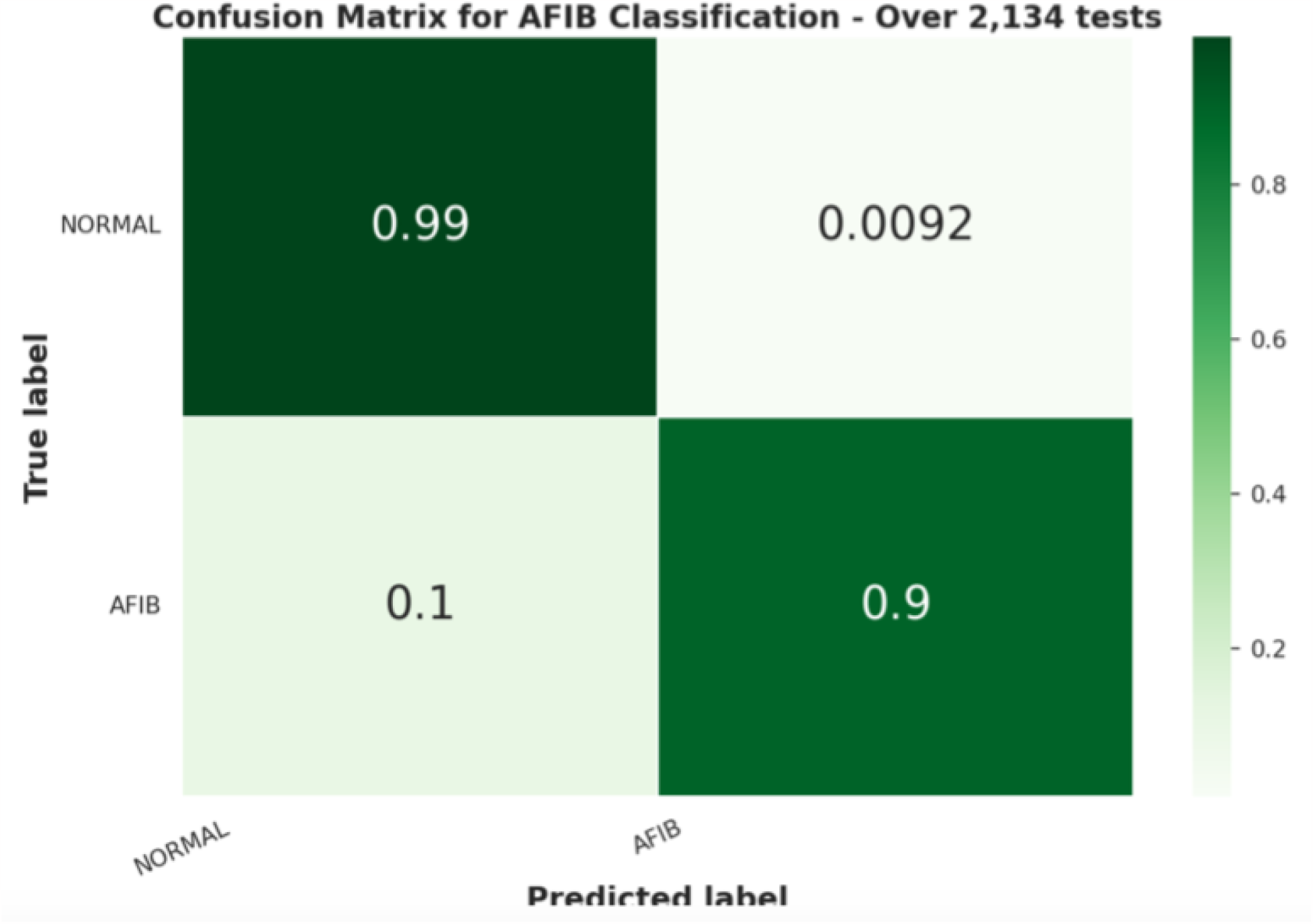
Confusion Matrix for ML method applied to MIMIC-III PPG dataset.

### H. Experiment 8 - UMMC

Secondly, the UMMC dataset was used to perform another round of tests on the trained model. Similarly, 30s-window-signals were extracted, processed, and evaluated using the two-step signal quality check. Figure 8 shows the dataset distribution. Table 6 and Figure14 show the results.

**Figure 14:**
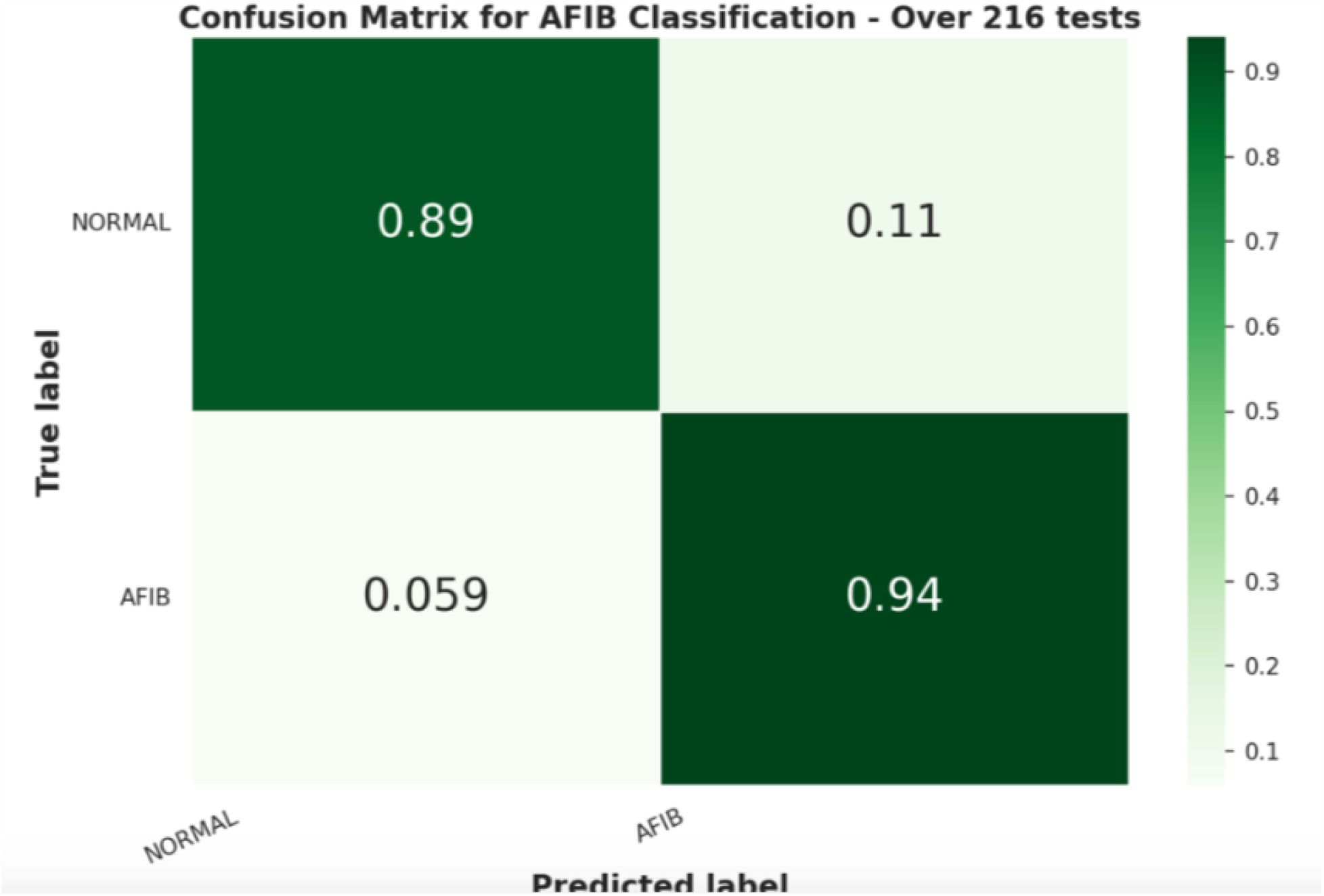
Confusion Matrix for ML method applied to UMMC PPG dataset.

**Table 6:** Statistical results for RF ML model trained using AFDB dataset and tested on UMMC dataset.

| Model | Acc. % | Prec. % | Recall % | F1-Score % |
| --- | --- | --- | --- | --- |
| RF | 89.81 | 80.15 | 91.56 | 84.03 |

Ultimately, Table 7 summarizes the RF model’s results across all five datasets.

**Table 7.** Statistical results for the ML method in all five datasets.

| Dataset | Acc. % | Prec. % | Recall % | F1-Score % |
| --- | --- | --- | --- | --- |
| AFDB | 96.62 | 96.62 | 96.62 | 96.62 |
| AFC | 92.64 | 80.08 | 86.06 | 82.68 |
| NSRDB | 92.86 | - | - | - |
| MIMIC-III | 97.32 | 96.68 | 94.37 | 95.47 |
| UMMC | 89.81 | 80.15 | 91.56 | 84.03 |

## DISCUSSION

The proposed methods show robust and reliable results across three different datasets.

The DSP method does not need a training process, however, it does need an optimal threshold to be chosen to correctly identify and avoid noisy parts of a signal.

In the first experiment, as can be seen in Figure 6, the DSP method was able to achieve a F1-score of 90.81% with 95% of predictions correctly identified as AFIB and 13% as False Positives, which shows a high rate of reliability on the model.

Moreover, the DSP method was tested on another dataset, as shown in Figure 7 Even though the performance decreased in comparison with previous results, it still remains acceptable, with 85% of predictions correctly identified as AFIB, which means that in 15% of the cases, the model missed an AFIB event. However, the false positive rate remained with 12 % of the Normal predictions being incorrectly classified as AFIB.

Lastly, in the NSRDB dataset, which generally exhibits good quality signals with Normal sinus rhythm data, the model was able to achieve 92.30% of accuracy (12 correct predictions out of 13) and represents that the method is robust in detecting Normal samples.

In the ML method, the datasets were highly augmented using HRV features extracted from individual frames, as can be seen in Figure 8. Following the extraction stage, the AFDB dataset had about 230,000 samples with almost 2 times more Normal samples than AFIB. For the other datasets, we ended up with 31,000 samples from the AFC dataset and 116,000 from the NSRDB dataset.

Unlike the DSP method, the ML method does require a training process, and 60% of the data from the AFDB dataset was used to train the model. However, those 60% were from individual samples to ensure that multiple samples from each individual don’t end up in both the training and test set. Three different ML models were compared in this work and were evaluated on the remaining 40% of the data. Results described in experiment 4 show that the random forest model performed the best, with a F1-Score of 95.67%. Moreover, Figure 9 shows that the RF model achieved a higher performance on the test set with 98% of the samples correctly identified AFIB predictions.

Furthermore, these results show the ability of the model in predicting frame-by-frame or in other words, for every 15 second window. This behavior is interesting given its applicability to real-time applications. However, sometimes utilizing a vote system for the final outcome is more reliable. Thus, the frames’ predictions were grouped by ID and a vote system was used to decide the final outcome. Across 114 samples (97.4%) the RF model correctly identified the non-AFIB diagnosis with only 3 errors, while 116 AFIB samples were correctly identified (96.7%) with only 4 errors. These results show that the model had success in detecting AFIB reliably across the test set.

However, in order to corroborate the results, the trained models were tested on a completely unseen dataset, and without additional training. In the AFC dataset, the SVM model performed slightly better as shown in Table 3, however this work chose to continue the analysis with the RF model due to the performance in further tests as well as the characteristics of an ensemble method. The RF model was able to achieve a F1-Score of 83%, which represents, as described in Figure 11, 95% of the Normal and 78% of the AFIB predictions correctly identified. Given the decrease in performance, we decided to apply a similar vote method, as we did in the previous test. As can be seen in Figure 12, the performance was almost the same as the frame predictions, for Normal and AFIB predictions, where there were 3,629 correctly identified Normal samples with 214 errors and 361 correct AFIB predictions with 105 errors.

Finally, in the NSRDB dataset, the RF model was able to achieve almost 94% accuracy in predicting Normal ECG signals in the frame-to-frame experiment. Moreover, when predicting the final outcome the model was able to predict correctly 13 out of 14 samples (92.86%).

All the results from the three datasets were combined in Table 4, showing that the RF model was able to achieve considerable results in detecting AFIB events as well as Normal samples, with a F1-score greater than 80% for all the datasets.

Moreover, experiments 7 and 8 describe the trained RF model tested on two other datasets which consisted of PPG signals only. Using a subset of the MIMIC-III dataset as the unseen test set, the RF model was able to achieve a F1-Score of 95%, where 99% of the Normal and 90% of AFIB predictions were correctly classified as described in Figure13.

Similarly, using the UMMC dataset the trained RF model was able to achieve a F1-Score of 84%, with 89% of Normal and 94% of AFIB predictions correctly identified. These results show that the ECG-based trained model was able to correctly identify AFIB events in PPG signals, moreover, this has shown robustness against false positives.

Ultimately, this work proposes a comparison with other authors that in their work, tried to achieve the same goal, AFIB detection using non-invasive real time techniques. Table 8 brings a comparison with three other authors, each one with a different approach, however all of them use a single dataset, AFDB.

**Table 8:** Performance of Classification Methods for Atrial Fibrillation Detection.

| **Authors** | **Accuracy %** | **Recall %** |
| --- | --- | --- |
| This work - DSP | 90.82 | 91.09 |
| This work - RF ML | 96.06 | 96.38 |
| Ross-Howe et al.<br>[7] | 93.16 | 98.33 |
| Bruun et al. [9] | 93.33 | 87.97 |
| Hu et al. [10] | 96.3 | 88.7 |

Through the comparison shown in Table 8, can be seen in the Recall metric that this work, using both DSP and the ML model, had slightly better performance than two authors, [9] and [10], while the approach proposed by Ross-Howe et al. [7] achieved the best performance with 98.33% of recall.

## CONCLUSION

In this paper, we propose two different methods for Atrial Fibrillation detection using short-term ECG signals. The first method uses only signal processing techniques to identify abrupt changes in the first derivative of the IBI signal. Using the ratio of abrupt changes with the number of peaks in a given frame, the DSP method classifies the condition using an optimal threshold.

In contrast with the DSP method, ML models were used. Seven HRV features were extracted from a 15 second ECG window and were used as input for different ML models.

Several experiments were conducted using three different ECG datasets that are publicly available. In the DSP method, all the data was used for testing, since there is not training process. Whilst, the ML models were trained using 60% of the individuals available in the AFDB dataset.

Results have shown that the DSP method was able to achieve 90.82%, 87.32% and 92.30% accuracy, respectively in the AFDB, AFC and NSRDB datasets. Similarly, the RF ML model was able to achieve 96.62%, 92.94% and 92.86% accuracy. The ECG-based trained RF ML model was chosen as the final solution and tested on two PPG datasets, without additional training.

Reported results show F1-Scores of 95.47% and 84% for MIMIC-III and UMMC dataset, respectively.

Ultimately, this work proposed a comparison with other authors that used the AFDB dataset to predict AFIB. This work showed slightly better performance when compared with two of them.

Even though this work has certain limitations against poor signal quality and noise influence, the benefits of identifying Atrial Fibrillation events in short-term ECG and PPG signals are a major breakthrough in healthcare research and large-scale monitoring.

## Data Availability

Researchers interested in access to the data may contact Nikhil Sehgal at, also see https://vastmindz.com/contact/. The authors will assist with any negotiable data use agreements and gain access to the data for any replication efforts following publication.

## COMPETING INTEREST

None

## ETHICS APPROVAL

Not Applicable

## FUNDING

None

## Notes

### Competing Interest Statement

The authors have declared no competing interest.

